# Potential spreading risks and disinfection challenges of medical wastewater by the presence of Severe Acute Respiratory Syndrome Coronavirus 2 (SARS-CoV-2) viral RNA in septic tanks of fangcang hospital

**DOI:** 10.1101/2020.04.28.20083832

**Authors:** Dayi Zhang, Haibo Ling, Xia Huang, Jing Li, Weiwei Li, Chuan Yi, Ting Zhang, Yongzhong Jiang, Yuning He, Songqiang Deng, Xian Zhang, Yi Liu, Guanghe Li, Jiuhui Qu

## Abstract

The outbreak of coronavirus infectious disease-2019 (COVID-19) pneumonia raises the concerns of effective deactivation of the severe acute respiratory syndrome coronavirus 2 (SARS-CoV-2) in medical wastewater by disinfectants. In this study, we evaluated the presence of SARS-CoV-2 viral RNA in septic tanks of Wuchang Fangcang Hospital and found the high level of (0.05-1.87)×10^4^ copies/L after disinfection with sodium hypochlorite. Embedded viruses in stool particles might be released in septic tanks, behaving as a source of SARS-CoV-2 and potentially contributing to its spread through drainage pipelines. Current recommended disinfection strategy (free chlorine above 6.5 mg/L after 1.5-hour contact) needs to be reevaluated to completely remove SARS-CoV-2 viral RNA in non-centralized disinfection system and effectively deactivate SARS-CoV-2. The effluents showed negative results for SARS-CoV-2 viral RNA when overdosed with sodium hypochlorite but had high a level of disinfection by-product residuals, possessing significant ecological risks.

## 1. Introduction

The outbreak of coronavirus infectious disease-2019 (COVID-19) pneumonia since 2019 is caused by the severe acute respiratory syndrome coronavirus 2 (SARS-CoV-2) (Lai et al., 2020; Li et al., 2020; Ralph et al., 2020) and it has rapidly spread throughout 202 countries around the world. Till 8^th^ May 2020, there are over 3.5 million confirmed cases and nearly 300,000 deaths globally, and the number is still increasing rapidly. There is clear evidence of human-to-human transmission of SARS-CoV-2 (Chan et al., 2020; Chang et al., 2020; Li et al., 2020; Poon and Peiris, 2020). Besides direct contact and respiratory routes (Carlos et al., 2020; Lai et al., 2020; Wu et al., 2020), stool transmission might be an alternative route owing to the presence and survival of SARS-CoV-2 in patient’s stools (Holshue et al., 2020; Ling et al., 2020; Tian et al., 2020; Xiao et al., 2020; Xing et al., 2020; Zhang et al., 2020). As municipal wastewater pipe network receives huge amounts of wastewater from residents and treated sewage from hospitals, SARS-CoV-2 from non- or inefficient-disinfected wastewater can persist for a prolonged time in pipe network, becoming a secondary spreading source. It brings urgent requirement and careful consideration of disinfection strategies to prevent SARS-CoV-2 from entering drainage pipe network.

Disinfection is of great importance to eliminate or deactivate pathogenic microorganisms. Traditional disinfection strategies include ultraviolet germicidal irradiation and biocidal agents, e.g., gaseous ozone, alcohol, formaldehyde, hydrogen peroxide, peroxyacetic acid, povidone iodine and chlorine-based disinfectant (Kampf et al., 2020; Tseng and Li, 2008; Tseng and Li, 2007; Walker and Ko, 2007). Chlorine-based disinfectants are widely used for their broad sterilization spectrum, high inactivation efficiency, low price, and easy decomposition with little residue (How et al., 2017). Nevertheless, overuse of chlorine-based disinfectants brings concerns of disinfection by-products (DBPs) which are harmful to ecosystems and human health (Bull et al., 2011; Richardson et al., 2007; Wang et al., 2014). More than 600 kinds of DBPs have been observed (Richardson, 2011), such as trihalomethanes (THMs), haloacetic acids (HAAs), halogen acetonitriles (HANs), halonitromethanes (HNMs) and haloacetamides (HAcAms) (Ding et al., 2020; Kozari et al., 2020; Luo et al., 2020; Zhai et al., 2014). For effective centralized disinfection, World Health Organization (WHO) has suggested free chlorine ≥ 0.5 mg/L after at least 30 minutes of contact time at pH<8.0 (WHO, 2020). Additionally, China has launched a guideline for emergency treatment of medical sewage containing SARS-CoV-2 on 1^st^ February 2020, requiring free chlorine of ≥ 6.5 mg/L and contact time of ≥1.5 hour in disinfection units (China-MEE, 2020). Unfortunately, the performance of chlorine-based disinfectants on SARS-CoV-2 in real medical wastewater treatment system is not clear yet.

In this work, we studied the presence of SARS-CoV-2 viral RNA in septic tanks of Wuchang Fangcang Hospital (Wuhan, China) to evaluate the disinfection performance and optimize disinfection strategies to prevent SARS-CoV-2 from spreading through drainage pipelines. Further analysis of DBPs evaluated the potential ecological risks in the effluents.

## 2. Materials and methods

### 2.1 Wuchang Fangcang Hospital and disinfection strategy

Wuchang Fangcang Hospital was open from 5^th^ February to 10^th^ March 2020, receiving 1124 COVID-19 patients (Figure 1). It had eight separate toilets and all sewage from toilets and showers were combined and disinfected in 4 preliminary disinfection tanks. They were then pumped into three septic tanks outside the hospital, followed a final disinfection (800 g/m^3^ of sodium hypochlorite before March 5^th^). After 24-hour, the effluent was pumped and discharged into pipe network and wastewater treatment plants. After 5 ^th^ March, the dosage of sodium hypochlorite was increased to 6700 g/m^3^ to secure complete deactivation of SARS-CoV-2.

**Figure 1.**
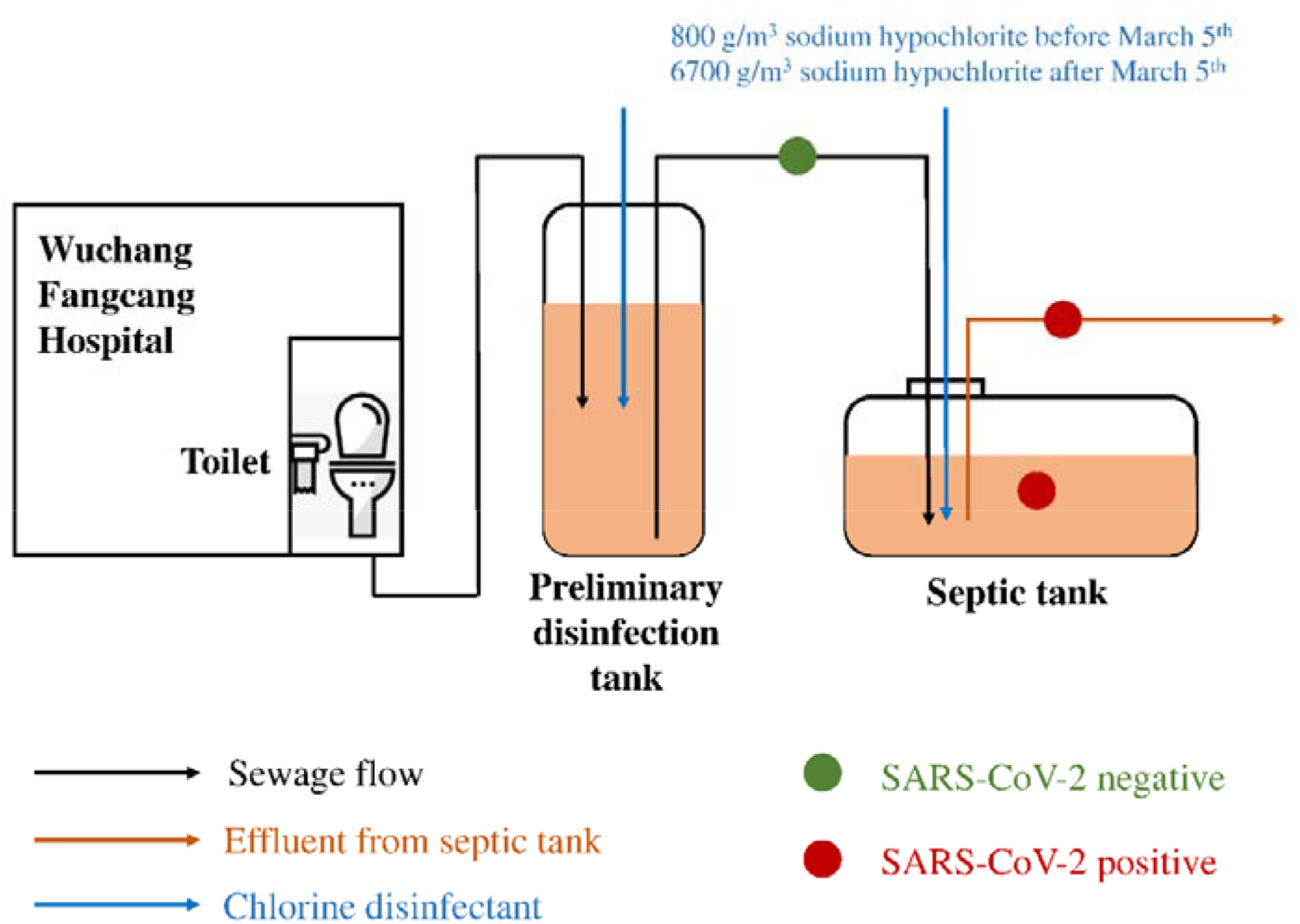
Schematic disinfection process of septic tanks of Wuchang Fangcang Hospital.

### 2.2 Sampling and chemical analysis

Influent and effluent samples were collected from septic tanks of Wuchang Fangcang Hospital on 26^th^ February, 1^st^ March and 10^th^ March, 2020. Around 2.0 L of water was directly collected in a plexiglass sampler and transferred into a sterile plastic bag for biological analysis and a brown glass bottle for DBPs analysis. A stratified plexiglass sampler was used to obtain samples from different layers of septic tanks, designated as top-layer (0-50 cm) and bottom-layer (50-100 cm) water.

Free chlorine was measured on site using PCII58700-00 (HACH, USA). DBPs measurements were carried out on a GCMS-QP2020 (Shimadzu, Japan) equipped with Atomx purge and trap autosampler (Teledyne Tekmar, USA). The autosampler operating conditions were as follows: purge for 11 min at 30 °C with high-purity nitrogen gas at a flow rate of 40 mL/min, dry purge for 1.0 min at 20 °C with the flow rate of 40 mL/min, pre-desorption at 180 °C and desorption for 2.0 min at 190 °C, and bake for 6.0 min at 200 °C.

### 2.3 RNA extraction and RT-qPCR

Collected water samples were placed in 4 °C ice-boxes and immediately transferred into laboratory for RNA extraction. After centrifugation at 3,000 rpm to remove suspended solids, the supernatant was subsequently supplemented with NaCl (0.3 mol/L) and PEG-6000 (10%), settled overnight at 4 °C, and centrifuged at 10,000 g for 30 minutes. Viral RNA in pellets was extracted using the EZ1 virus Mini kit (Qiagen, Germany) according to the manufacturer’s instructions. SARS-CoV-2 RNA was quantified by RT-qPCR using AgPath-ID™ One-Step RT-PCR Kit (Life Technologies, USA) on a LightCycler 480 Real-time PCR platform (Roche, USA) in duplicates. Two target genes simultaneously amplified were open reading frame lab (CCDC-ORF1, forwards primer: 5'-CCCTGTGGGTTTTACACTTAA-3'; reverse primer: 5'-ACGATTGTGCATCAGCTGA-3'; fluorescence probe: 5 '-FAM-C CGTCTGCGGTATGTGGAAAGGTTATGG-BHQ1-3') and nucleocapsid protein (CCDC-N, forwards primer: 5'-GGGGAACTTCTCCTGCTAGAAT-3'; reverse primer: 5'-CAGACATTTTGCTCTCAAGCTG-3'; fluorescence probe: 5'-FAM-TTGCTGCTGCTTGACAGATT-TAMRA-3'). RT-qPCR amplification for CCDC-ORF1 and CCDC-N was performed in 25 μL reaction mixtures containing 12.5 μL of 2xRT-PCR Buffer, 1 μL of 2×RT-PCR Enzyme Mix, 4 μL mixtures of forward primer (400 nM), reverse primer (400 nM) and probe (120 nM), and 5 μL of template RNA. Reverse transcription was conducted at 45 °C for 10 min (1 cycle), followed by initial denaturation at 90 °C for 10 min (1 cycle) and 40 thermal cycles of 60 °C for 45 second and 90 °C for 15 seconds. Quantitative fluorescent signal for each sample was normalized by ROX™ passive reference dye provided in 2xRT-PCR buffer. For each RT-qPCR run, both positive and negative controls were included. The copy numbers of SARS-CoV-2 was obtained from a standard calibration curve by a 10-fold serial dilution of genes encoding nucleocapsid protein with an amplification efficiency of 102.6%, calculated as copies=10^[-(Cq-39086)/3 262]^ (R^2^=0.991). For quality control, a reagent blank and extraction blank were included for RNA extraction procedure and no contamination was observed.

### 2.4 Data analysis

One-way ANOVA was used to compare the difference between samples and p-value less than 0.05 refers to significant difference.

## 3. Results and discussions

For all influents of septic tanks received from Wuchang Fangcang Hospital, there was no positive result for SARS-CoV-2 viral RNA. On 26^th^ February and 1^st^ March, SARS-CoV-2 viral RNA was 1.47×10^4^ and (0.05-1.87)×10^4^ copies/L in the effluents of septic tanks, respectively (Table 1). Although there was no significant difference between these two sampling days, there were more SARS-CoV-2 RNA in top-layer waters (averagely 1.00× 10^4^ copies/L) than in bottom-layer waters (averagely 0.51×10^4^ copies/L, *p*=0.019).

**Table 1.**
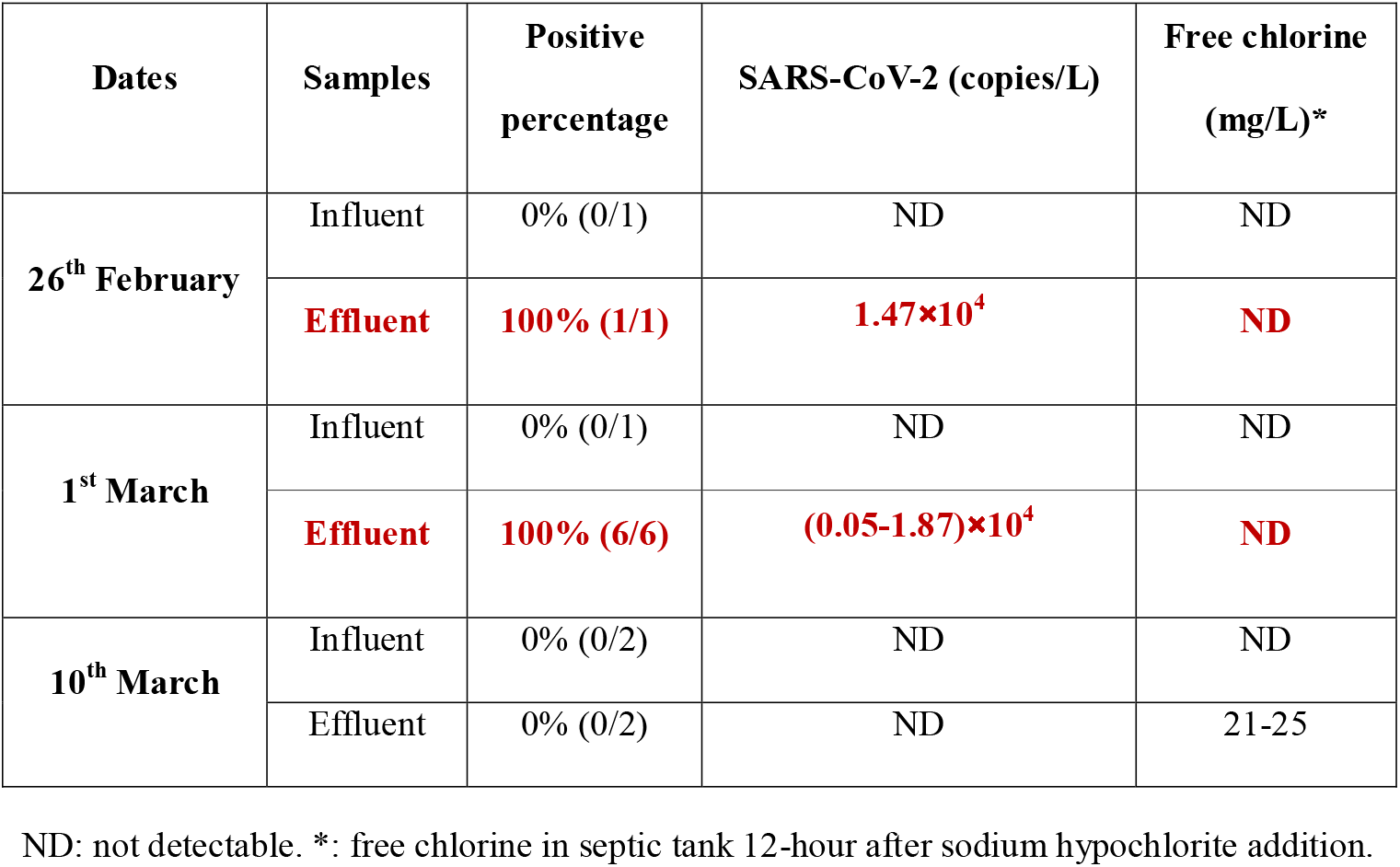
Copy numbers of SARS-CoV-2 viral RNA and free chlorine in the effluents of septic tanks of Wuchang Fangcang Hospital.

| Dates | Samples | Positive percentage | SARS-CoV-2 (copies/L) | Free chlorine (mg/L)* |
| --- | --- | --- | --- | --- |
| 26<sup>th</sup> February | Influent | 0% (0/1) | ND | ND |
|  | **Effluent** | **100% (1/1)** | **1.47×10<sup>4</sup>** | **ND** |
| 1<sup>st</sup> March | Influent | 0% (0/1) | ND | ND |
|  | **Effluent** | **100% (6/6)** | **(0.05-1.87)×10<sup>4</sup>** | **ND** |
| 10<sup>th</sup> March | Influent | 0% (0/2) | ND | ND |
|  | Effluent | 0% (0/2) | ND | 21-25 |

ND: not detectable. \*: free chlorine in septic tank 12-hour after sodium hypochlorite addition.

On 26^th^ February and 1^st^ March, free chlorine in the effluents was above 6.5 mg/L after 1.5 hours contact with 800 g/m^3^ of sodium hypochlorite, meeting well with the guideline for emergency treatment of medical sewage containing SARS-CoV-2 suggested by China CDC. Twelve hours after sodium hypochlorite addition, free chlorine declined to nondetectable (total chlorine 4.0-8.8 mg/L) when SARS-CoV-2 viral RNA were detectable (Table 1). It hinted an unexpected presence of SARS-CoV-2 viral RNA after disinfection and a dosage of 800 g/m^3^ of sodium hypochlorite could not completely remove SARS-CoV-2 viral RNA. To improve disinfection performance and secure a complete deactivation of SARS-CoV-2, the dosage of sodium hypochlorite was increased to 6700 g/m^3^ since 6^th^ March and free chlorine in the effluents on 10^th^ March ranged from 21.0 to 25.0 mg/L at 12-hour after sodium hypochlorite addition. Eventually, both influents and effluents of septic tanks were negative for SARS-CoV-2 viral RNA. However, trichlormethane, tribromomethane, bromodichloromethane and dibromochloromethane was 182-482, 0.6-3.1, 1.3-8.9 and ND-1.2 μg/L in the effluents, respectively.

Our results suggested that disinfection in Wuchang Fangcang Hospital was satisfactory to deactivate SARS-CoV-2 in aqueous phase after 1.5-hour contact, owing to the absence of SARS-CoV-2 viral RNA. Addition of 800 g/m^3^ of sodium hypochlorite according to the guideline for emergency treatment of medical sewage containing SARS-CoV-2 suggested by China CDC achieved free chlorine > 6.5 mg/L for 1.5 hours, but SARS-CoV-2 viral RNA was present in the effluents at 12-hour after sodium hypochlorite addition when free chlorine declined to nondetectable. SARS-CoV-2 in patients’ stools might explain the surprising presence of SARS-CoV-2 viral RNA in the effluents (Wu et al., 2020; Xing et al., 2020), that viruses embedded in stool particles could escape from disinfection and slowly release into aqueous phase. Suspended particles as small as 7 mm can protect viruses from UV exposure and dwindle their vulnerability to direct sunlight inactivation, and 0.3-mm sized particles can shield viruses from disinfection for their prolonged survival (Templeton et al., 2005). As stools are rich in organic compounds and form numerous suspended solids containing SARS-CoV-2, they are of high risk as the source releasing viruses in septic tanks. It is also evidenced by fewer SARS-CoV-2 RNA copy numbers in upper-layer waters, explained by more stool residuals and suspended solids in the bottom of septic tanks absorbing SARS-CoV-2 from aquatic water. Thus, septic tanks might behave as a long-term source to release SARS-CoV-2 viral RNA into waters when disinfection is not completed and challenges public health *via* potentially spreading viruses in drainage pipelines.

The surprising presence of SARS-CoV-2 viral RNA after disinfection with sodium hypochlorite suggested that free chlorine > 0.5 mg/L after 1.5-hour contact time is not efficient for completely removing SARS-CoV-2 viral RNA, and 800 g/m^3^ dosage of sodium hypochlorite might be not enough to secure a complete disinfection of medical wastewaters, particularly for those from fangcang, temporary or non-centralized hospitals. From the negative results of SARS-CoV-2 viral RNA (Table 1), the complete deactivation of SARS-CoV-2 was achieved when the dosage of sodium hypochlorite was 6700 g/m^3^. Nevertheless, it was an over-dosage and resulted in a significantly higher level of DPB residues in the effluents than other reports (Luo et al., 2020). They show high ecological risks and challenge the surrounding environment receiving disinfected medical wastewater. Additionally, applying high level of chlorine-based and other disinfectants have lasted for three months in Wuhan since the outbreak of COVID-19, and further studies are suggested to carefully evaluate its ecological risks.

Owing to the operational limitations during the COVID-19 outbreak in Wuhan and restricted sample transport to other laboratories, this study did not demonstrate viral viability after disinfection by viral culture. Additionally, disinfection operations in septic tanks of Wuchang Fangcang Hospital were strictly controlled by government officials that we could not collect more samples or explore the optimal dosage of sodium hypochlorite for complete removal of SARS-CoV-2 viral RNA with minimal DBPs. This work attempts to give initial information about the potential risks of viral spread and DBPs residues during disinfection processes for medical wastewater containing SARS-CoV-2, and further studies are required to confirm these preliminary results.

## 4. Conclusion

Our study for the first time reported an unexpected presence of SARS-CoV-2 viral RNA in septic tanks after disinfection with 800 g/m^3^ of sodium hypochlorite and current disinfection guideline by WHO and China CDC might not secure a complete removal of SARS-CoV-2 in medical wastewater. SARS-CoV-2 might be embedded in patient’s stools, protected by organic matters from disinfection, and slowly release when free chlorine declines. Septic tanks in non-centralized disinfection system of fangcang hospitals or isolation points potentially behave as a secondary source spreading SARS-CoV-2 in drainage pipelines for prolonged time. Disinfection strategy is of great urgency to improve and the ecological risks of DBPs need to be carefully considered.

## Data Availability

All data used in the study are available from the corresponding author by request.

## 5. Author contributions

Concept and design: DZ, XH, YL, JQ.

Acquisition, analysis, or interpretation of data: DZ, HL, JL, WL, CY, TZ, YJ, XZ, GL.

Drafting of the manuscript: DZ, YH, SD.

Statistical analysis: DZ.

